# The effect of circulating zinc, selenium, copper and vitamin K_1_ on COVID-19 outcomes: a Mendelian randomization study

**DOI:** 10.1101/2021.10.18.21265128

**Authors:** Maria K. Sobczyk, Tom R. Gaunt

## Abstract

**Background:** Previous results from observational, interventional studies and in vitro experiments suggest that certain micronutrients have anti-viral and immunomodulatory activities. In particular, it has been hypothesized that zinc, selenium, copper and vitamin K_1_ have strong potential for prophylaxis and treatment of COVID-19.

**Objectives:** We aimed to test whether genetically predicted Zn, Se, Cu or vitamin K_1_ levels have a causal effect on COVID-19 related outcomes: risk of infection, hospitalization and critical illness.

**Methods:** We employed two-sample Mendelian Randomization (MR) analysis. Our genetic variants derived from European-ancestry GWAS reflected circulating levels of Zn, Cu, Se in red blood cells as well as Se and vitamin K_1_ in serum/plasma. For the COVID-19 outcome GWAS, we used infection, hospitalization or critical illness. Our inverse-variance weighted (IVW) MR analysis was complemented by sensitivity analyses: more liberal selection of variants at genome-wide subsignificant threshold, MR-Egger and weighted median/mode tests.

**Results:** Circulating micronutrient levels show limited evidence of association with COVID-19 infection with odds ratio [OR] ranging from 0.97 (95% CI: 0.87-1.08, *p*-value=0.55) for zinc to 1.07 (95% CI: 1.00-1.14, *p*-value=0.06) – ie. no beneficial effect for copper, per 1 SD increase in exposure. Similarly minimal evidence was obtained for the hospitalization and critical illness outcomes with OR from 0.98 (95% CI: 0.87-1.09, *p*-value=0.66) for vitamin K1 to 1.07 (95% CI: 0.88-1.29, *p*-value=0.49) for copper, and from 0.93 (95% CI: 0.72-1.19, *p*-value=0.55) for vitamin K_1_ to 1.21 (95% CI: 0.79-1.86, *p*-value=0.39) for zinc, respectively.

**Conclusions:** This study does not provide evidence that supplementation with zinc, selenium, copper or vitamin K_1_ can prevent SARS-CoV-2 infection, critical illness or hospitalization for COVID-19.

## Introduction

Highly transmissible and virulent in at-risk groups, SARS-CoV-2, the causal agent for COVID-19, has been sweeping the globe since December 2019. However, despite intensive research, there are few effective prophylactic and early-stage therapeutic interventions for COVID-19, with the exception of vaccines(1,2). However, worldwide vaccine distribution remains highly inequitable, with less than 4% of the African population vaccinated as of September 2021(3).

Corollary to drug repurposing efforts, the potential role of micronutrient supplementation towards preventing and alleviating COVID-19 has been proposed(4,5). Supplements have some unique advantages as they are inexpensive, widely available over-the-counter, easily distributed and stored, generally well tolerated and well characterised in terms of safety. Amongst vitamins and minerals, good mechanistic reasons for more research exist for zinc, selenium, copper and vitamin K_1_, chiefly due to their important roles in immune and antiviral response(4).

Zinc (Zn) is an essential trace metal with structural roles in regulatory proteins, as enzyme cofactor, and as a signalling molecule. Of relevance to COVID-19, zinc deficiency can lead to dysfunctional immune response with reduced activity of innate immune cells(6,7), lymphopenia(8) and activation of NF-KB signalling inducing production of IL-6 and other cytokines involved in “cytokine storm”, characteristic of COVID-19(9–11). Furthermore, zinc has manifold antiviral properties in vitro and in vivo(12,13). With respect to SARS-CoV, Zn combined with a ionophore was shown to inhibit its RNA polymerase and block virus replication in cell culture(14). In SARS-CoV-2, Zn^2+^ inhibits the main protease (M^pro^) which results in reduced viral replication in cell culture(15). Limited evidence from randomized trials on common cold suggests beneficial effect of zinc supplementation on cold duration and less conclusively, incidence and severity(16), while adjuvant treatment in severe paediatric pneumonia reduced hospital stay(17).

Selenium (Se) is a constituent of 25 selenoproteins with functions in redox homeostasis, endoplasmic reticulum stress and inflammatory response(18). Overall, Se can be a partial determinant of viral virulence(19). Sub-optimally low Se intake is combined with coxsackievirus infection in aetiology of Keshan disease(20). Furthermore, immunocompetence for infection clearance with other viral diseases is decreased in Se deficiency(21) and marginal Se status(22). In vitro, Se supplementation was shown to inhibit replication of porcine circovirus(23). Ebselen, a synthetic organoselenium compound, was found to be one of the most effective SARS-CoV-2 main protease (M^pro^) inhibitors by forming a selenyl sulphide bond with the protease’s catalytic dyad(24,25) which provides a potential mode of antiviral action for organic selenium molecules, similarly to ionic zinc. In addition, ebselen is functionally related to glutathione peroxidase 1, a major selenoenzyme which has been also found to physically interact with M^pro^(26). Other mechanisms through which Se could help in COVID-19 management is through control of ROS-driven endothelial damage(18), reduced IL-6 pathway response(27–29) and stimulation of innate immune system(30,31).

Copper (Cu) is indispensable for the processes of respiration, free radical defence and immune regulation due to its structural role in cuproenzymes(32,33). Similar to Zn, Cu plays an essential part in antioxidant response activated during inflammation. Copper deficiency can result in neutropenia and immunosuppression via reduced T-cell proliferation(34). Inactivation of viruses, including SARS-CoV-2(35) on copper surfaces is widely exploited in clinical practice(36), but Cu^2+^ was also reported to decrease infectivity of HIV(37), influenza(38) virus and SARS-CoV-2(39) in mammalian cells. Copper can also exert antiviral properties potentially by stimulating autophagy(40).

Two vitamers of vitamin K exist: K_1_ (phylloquinone) and K_2_ (menaquinone)(41). Vitamin K is necessary for activation of pro- and anti-clotting factors in the liver and peripheral tissues, respectively. Moreover, vitamin K activates Matrix Gla protein (MGP) which inhibits elastic fibre degradation and vascular mineralisation. Extrahepatic vitamin K deficiency and low MGP activity have been found in hospitalised COVID-19 patients(42,43). According to Janssen et al (2021)(44), this could result from increased degradation of elastic fibres by SARS-CoV-2 promoting lung fibrosis and concomitant with predicted depletion of endothelial vitamin K-dependent anti-coagulant (protein S) lead to coagulopathy. Therefore, vitamin K could provide adjunct therapy of thrombosis events which are characteristic of severe COVID-19(45).

In the absence of well-powered randomized control trials (RCT) testing the prophylactic and therapeutic potential of these micronutrients, we decided to carry out a Mendelian Randomization (MR) assessment of their potential causal effects. MR is an established causal inference method which uses genetic variants as instrumental variables(46).

Since genetic variants are independently and randomly distributed at meiosis and established at conception, the risk of confounding and reverse causation is greatly reduced in MR (46). This is especially important as any observational studies linking nutrients levels to COVID-19 outcomes will be confounded by fact that COVID-19 at-risk groups (e.g. the elderly, high BMI individuals, diabetics(47)) have on average lower/suboptimal levels of many micronutrients(8,10,13,42,44,48–50) and at the same time suffer from poorer COVID-19 outcomes. MR analyses can help to clarify the causal pathway in such cases. As such, MR has been successfully applied in nutritional epidemiology(51), including in studies using our micronutrients of interest as exposure(52–58).

## Methods

### Selection of genetic instruments – relevance MR criterion

#### GWAS studies

We searched the literature, OpenGWAS(59) and GWAS Catalog(60) for genetic instruments associated with zinc, copper, selenium and vitamin K_1_ levels in populations of European ancestry.

We evaluated genetic instruments from the published GWAS of zinc, copper and selenium content of erythrocytes in the Queensland Institute of Medical Research (QIMR) twin cohort and whole blood selenium in the Avon Longitudinal Study of Parents and Children (ALSPAC) cohort of pregnant women (61) measured using inductively coupled plasma mass spectrometry. Red blood concentrations of those trace elements generally represent overall nutritional status well(62) and total blood measurement is a standard biomarker(63). GWAS was adjusted for the following covariates: analysis batch, haemoglobin concentration and analytical QC data.

For selenium instruments, we used a published fixed-effects meta-analysis of toe-nail selenium concentration measured using neutron activation analysis in four European-ancestry US cohorts (Coronary Artery Risk Development in Young Adults, Johnston County Osteoarthritis Project, Nurses’ Health Study, Health Professionals Follow-up Study) co-analysed with the QIMR & ALSPAC GWAS results(64). Toe-nail Se GWAS was adjusted for the following covariates: age, smoking status, geography and top eigenvectors. Compared to circulating selenium, toe-nail content reflects more long-term Se exposure.

Vitamin K instruments were derived from a GWAS of phylloquinone (vitamin K_1_), the primary circulating form of vitamin K, which measurements were available in 2 European ancestry CHARGE cohorts(65): Framingham Offspring Consortium and the Health, Aging and Body Composition study. Phylloquinone measurements were taken in plasma/serum using reverse-phase high-performance liquid chromatography followed by fluorometric detection. Vitamin K_1_ concentration in plasma reflects recent intake(42). The GWAS included the following covariates: age, sex and study-specific stratification.

For each micronutrient phenotype, we clumped the instruments using PLINK ver. v1.90b4.1(66) and 1000 Genomes European reference panel(67), at the threshold of *r*^2^ < 0.05 and clumping distance of 10 Mbp. The final set of instruments was derived by combining 1 representative SNP from each clump selected based on lowest *p*-value and presence in the outcome dataset.

#### Zinc genetic instruments

One of the two instruments for zinc was missing (rs1532423), however we found a good proxy for it using 1000 Genomes European population using the LDproxy web app(68): rs2453868, situated 34,383 bp away with *r*^2^ of 0.93 and *D’* of 1 (correlated alleles: rs1532423_A_= rs2453868_C_, rs1532423_G_= rs2453868_T_). Altogether, rs2120019 and rs1532423 account for 4.6% of variance in red blood cell copper concentration (**Supplementary Table 1**).

#### Selenium genetic instruments

Out of 12 genome-wide significant SNPs found at 2 loci in the meta-analysis, 4 SNPs survived LD-pruning. However, only 2 variants could be used as instruments (rs921943, rs6859667) as SNPs in the other 2 clumps (rs6586282, rs1789953, rs234709) or their proxies were not available in the COVID-19 GWAS. The 2 instruments in the meta-analysis accounted for only 2.25% of variance in the trait (Supplementary Table 1). The results of the meta-analysis were initially presented as Z-scores which we converted to betas using the formula in Taylor et al. (2016)(69).

#### Copper genetic instruments

Two genome-wide significant instruments were identified: rs1175550 and rs2769264; altogether they account for 4.6% of variance in red blood cell copper concentration (Supplementary Table 1).

#### Vitamin K_1_ genetic instruments

11 signals at 6 loci were detected in the GWAS discovery stage at *p-*value < 5 × 10^−5^, with none of the SNPs reaching genome-wide significance. We found four SNPs of concern which were removed from downstream processing (**Supplementary Methods**). The three variants retained: rs4645543, rs4852146, rs6862071 (Supplementary Table 1) jointly explained 3.06% of variance in circulating phylloquinone concentration.

#### Sensitivity analyses using subsignificant hits

In addition to the main analysis where we included only genome-wide significant hits at *p*-value 5 × 10^−8^ (with the exception of vitamin K_1_, where no variants crossed the threshold), we ran sensitivity analyses including variants selected at a more liberal, subsignificant threshold of 5 × 10^−5^. This allowed us to increase statistical power of analysis at the increased risk of violation of core MR assumptions. For zinc (**Supplementary Table 2**), we used 12 SNPs (total R^2^ of 14%), while for selenium we only had access to results from individual cohorts: QIMR (15 SNPs, R^2^ = 13.26%) and ALSPAC (12 SNPs, R^2^ = 10.93%) and for copper we arrived at 7 SNPs (R^2^ = 10.03%).

### Independence MR criterion

We calculated the variance in each exposure explained by each set of instruments (*R*^2^) and F-statistics using the formulas in Yarmolinsky et al. (2018)(55). We did not see any weak instrument bias, with F-statistic ranging from 15 to 172 (Supplementary Table 1 and 2).

### Exclusion restriction MR criterion

PhenoScanner V2(70) was used to assess presence of horizontal pleiotropy among the candidate variants using default settings (**Supplementary Table 3** and **4**, Supplementary Methods).

In addition, we carried out leave-one-out analysis in our sensitivity checks which should minimize any possible confounding introduced by individual SNPs associated with height and RBC traits, whenever possible. Next, we calculated Cochran’s Q statistic and I^2^ to look for signs of heterogeneity, also indicative of pleiotropy. Finally, MR-Egger is one of the MR methods which we employed and which can detect directional horizontal pleiotropy if the intercept significantly deviates from 0(71).

### Selection of outcomes

The biggest publicly available GWAS to date on COVID-19 from the COVID-19 Host Genetics Initiative release 5 was chosen(72). This fixed-effect meta-analysis contains up to 49,562 COVID-19 patients and 2 million controls from 46 studies across 19 countries, however, we used results obtained in 35 European-only cohorts. The outcomes available were very severe (critical) COVID-19 (vs population), hospitalized (vs SARS-CoV-2 infected but non-hospitalized with COVID-19 or vs population) and SARS-CoV-2 infection (vs population) (**Supplementary Table 5**). The covariates used in the GWAS analysis were age and sex.

### Statistical analysis

We used the online mRnd power calculator to explore the limits of our MR analysis(73). All the MR analyses were done using TwoSampleMR(74) and MendelianRandomization(75) R packages. Our main method was the inverse variance weighted (IVW) random-effects meta-analysis of causal effects of individual instruments as it is the most efficient, however IVW is biased in cases of unbalanced pleiotropy(76). We complemented IVW with analyses using other MR methods: MR-Egger(71), weighted median-based and mode-based estimator. MR-Egger relaxes the assumption of balanced horizontal pleiotropy at the cost of reduced power, whereas weighted median estimator is still valid if only at least 50% of variants meet the three main MR assumptions(46); mode-based estimator is similarly robust to outliers while being more conservative.

### Ethics

This study uses publicly available summary data and no original data collection was undertaken for this manuscript. Evidence of ethical approval for all of the included GWAS studies can be found in the previous publications. Our investigation is in accordance with the ethical guidelines of the 1975 Declaration of Helsinki.

## Results

### Power analysis

In the main analysis, SARS-CoV-2 infection was the outcome with the biggest power due to highest number of cases and controls in the outcome GWAS. Minimum detectable one-tailed odds ratio (OR) at 80% power ranged from 0.91 for selenium to 0.93 for zinc and copper (**Supplementary Table 6)**. COVID-19 hospitalization analysis possessed lower power ranging from OR of 0.82 for Se to 0.87 for Zn and Cu. Poor power was found for very severe COVID-19 (OR of 0.75 for Se to 0.81 for Zn and Cu). In order to increase power in our sensitivity analyses, we used sets of genome-wide subsignificant (max *p-*value of 5 × 10^−5^) variants for zinc, selenium and copper (**Supplementary Table 7**). In the subsignificant instrument analyses, minimum detectable one-tailed odds ratios at 80% power were 0.95-0.96 for SARS-CoV-2 infection and 0.87-0.89 for very severe (critical) COVID-19.

### Mendelian Randomization

#### Zinc

We found little evidence of genetically-predicted circulating zinc concentration having a large effect on COVID-19 outcomes (**Table 1, Figure 1**). IVW odds ratios of SARS-CoV-2 infection were 0.97 (95% CI: 0.87-1.08, nominal *p*-value=0.55) in the main analysis and 1.01 (95% CI: 0.98-1.05, nominal *p*-value=0.49) in the sensitivity subsignificant (max *p-*value of 5 × 10^−5^) analysis, per 1 SD increase of circulating zinc. Hospitalization (ver. population) showed weak evidence of association with zinc levels (OR=1.06, 95% CI: 0.81-1.39, *p*-value=0.66) in the main analysis and subsignificant analysis (OR=0.98, 95% CI: 0.91-1.06, *p*-value=0.62). Similar results were obtained for the hospitalized (ver. non-hospitalized) outcome (Figure 1). Lastly, we did not find any strong effect of zinc on very severe disease in the main analysis (OR=1.21, 95% CI: 0.79-1.86, *p*-value=0.39). A narrower but overlapping estimate was derived from the more liberal set of SNPs (OR=0.92, 95% CI: 0.81-1.04, *p-*value=0.16). Estimates in the sensitivity analyses using MR-Egger, median-weighted and mode-weighted estimator directionally matched the results from IVW (Figure 1, **Supplementary Table 8**).

**Table 1.**
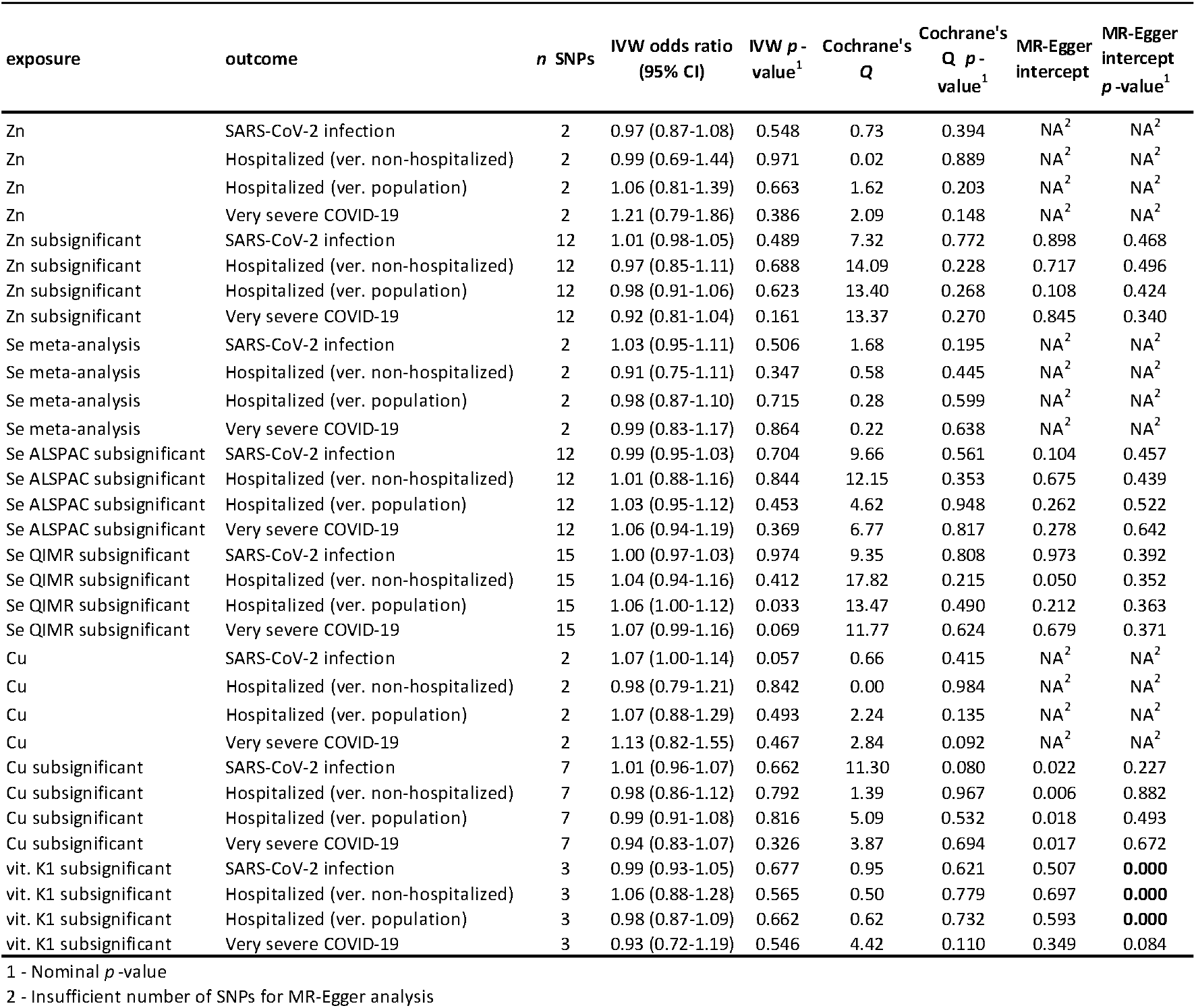
Results of MR analysis studying the effect of circulating zinc (Zn), selenium (Se), copper (Cu) and vitamin K_1_ concentration on 3 COVID-19 outcomes. Inverse-variance weighted (IVW)-based odds ratios, their *p*-values along with Cochrane’s Q statistic and MR-Egger intercept are presented. We used two sets of instruments whenever possible: Zn/Se/Cu refers to instruments with *p*-values < 5 × 10^−8^ and Zn/Se/Cu/vit. K_1_ subsignificant refers to instruments with *p*-values < 5 × 10^−5^.

| exposure | outcome | *n* SNPs | IVW odds ratio<br>(95% CI) | IVW *p*-<br>value<sup>1</sup> | Cochrane's<br>Q | Cochrane's<br>Q *p*-<br>value<sup>1</sup> | MR-Egger<br>intercept | MR-Egger<br>intercept<br>*p*-value<sup>1</sup> |
| --- | --- | --- | --- | --- | --- | --- | --- | --- |
| Zn | SARS-CoV-2 infection | 2 | 0.97 (0.87-1.08) | 0.548 | 0.73 | 0.394 | NA<sup>2</sup> | NA<sup>2</sup> |
| Zn | Hospitalized (ver. non-hospitalized) | 2 | 0.99 (0.69-1.44) | 0.971 | 0.02 | 0.889 | NA<sup>2</sup> | NA<sup>2</sup> |
| Zn | Hospitalized (ver. population) | 2 | 1.06 (0.81-1.39) | 0.663 | 1.62 | 0.203 | NA<sup>2</sup> | NA<sup>2</sup> |
| Zn | Very severe COVID-19 | 2 | 1.21 (0.79-1.86) | 0.386 | 2.09 | 0.148 | NA<sup>2</sup> | NA<sup>2</sup> |
| Zn subsignificant | SARS-CoV-2 infection | 12 | 1.01 (0.98-1.05) | 0.489 | 7.32 | 0.772 | 0.898 | 0.468 |
| Zn subsignificant | Hospitalized (ver. non-hospitalized) | 12 | 0.97 (0.85-1.11) | 0.688 | 14.09 | 0.228 | 0.717 | 0.496 |
| Zn subsignificant | Hospitalized (ver. population) | 12 | 0.98 (0.91-1.06) | 0.623 | 13.40 | 0.268 | 0.108 | 0.424 |
| Zn subsignificant | Very severe COVID-19 | 12 | 0.92 (0.81-1.04) | 0.161 | 13.37 | 0.270 | 0.845 | 0.340 |
| Se meta-analysis | SARS-CoV-2 infection | 2 | 1.03 (0.95-1.11) | 0.506 | 1.68 | 0.195 | NA<sup>2</sup> | NA<sup>2</sup> |
| Se meta-analysis | Hospitalized (ver. non-hospitalized) | 2 | 0.91 (0.75-1.11) | 0.347 | 0.58 | 0.445 | NA<sup>2</sup> | NA<sup>2</sup> |
| Se meta-analysis | Hospitalized (ver. population) | 2 | 0.98 (0.87-1.10) | 0.715 | 0.28 | 0.599 | NA<sup>2</sup> | NA<sup>2</sup> |
| Se meta-analysis | Very severe COVID-19 | 2 | 0.99 (0.83-1.17) | 0.864 | 0.22 | 0.638 | NA<sup>2</sup> | NA<sup>2</sup> |
| Se ALSPAC subsignificant | SARS-CoV-2 infection | 12 | 0.99 (0.95-1.03) | 0.704 | 9.66 | 0.561 | 0.104 | 0.457 |
| Se ALSPAC subsignificant | Hospitalized (ver. non-hospitalized) | 12 | 1.01 (0.88-1.16) | 0.844 | 12.15 | 0.353 | 0.675 | 0.439 |
| Se ALSPAC subsignificant | Hospitalized (ver. population) | 12 | 1.03 (0.95-1.12) | 0.453 | 4.62 | 0.948 | 0.262 | 0.522 |
| Se ALSPAC subsignificant | Very severe COVID-19 | 12 | 1.06 (0.94-1.19) | 0.369 | 6.77 | 0.817 | 0.278 | 0.642 |
| Se QIMR subsignificant | SARS-CoV-2 infection | 15 | 1.00 (0.97-1.03) | 0.974 | 9.35 | 0.808 | 0.973 | 0.392 |
| Se QIMR subsignificant | Hospitalized (ver. non-hospitalized) | 15 | 1.04 (0.94-1.16) | 0.412 | 17.82 | 0.215 | 0.050 | 0.352 |
| Se QIMR subsignificant | Hospitalized (ver. population) | 15 | 1.06 (1.00-1.12) | 0.033 | 13.47 | 0.490 | 0.212 | 0.363 |
| Se QIMR subsignificant | Very severe COVID-19 | 15 | 1.07 (0.99-1.16) | 0.069 | 11.77 | 0.624 | 0.679 | 0.371 |
| Cu | SARS-CoV-2 infection | 2 | 1.07 (1.00-1.14) | 0.057 | 0.66 | 0.415 | NA<sup>2</sup> | NA<sup>2</sup> |
| Cu | Hospitalized (ver. non-hospitalized) | 2 | 0.98 (0.79-1.21) | 0.842 | 0.00 | 0.984 | NA<sup>2</sup> | NA<sup>2</sup> |
| Cu | Hospitalized (ver. population) | 2 | 1.07 (0.88-1.29) | 0.493 | 2.24 | 0.135 | NA<sup>2</sup> | NA<sup>2</sup> |
| Cu | Very severe COVID-19 | 2 | 1.13 (0.82-1.55) | 0.467 | 2.84 | 0.092 | NA<sup>2</sup> | NA<sup>2</sup> |
| Cu subsignificant | SARS-CoV-2 infection | 7 | 1.01 (0.96-1.07) | 0.662 | 11.30 | 0.080 | 0.022 | 0.227 |
| Cu subsignificant | Hospitalized (ver. non-hospitalized) | 7 | 0.98 (0.86-1.12) | 0.792 | 1.39 | 0.967 | 0.006 | 0.882 |
| Cu subsignificant | Hospitalized (ver. population) | 7 | 0.99 (0.91-1.08) | 0.816 | 5.09 | 0.532 | 0.018 | 0.493 |
| Cu subsignificant | Very severe COVID-19 | 7 | 0.94 (0.83-1.07) | 0.326 | 3.87 | 0.694 | 0.017 | 0.672 |
| vit. K1 subsignificant | SARS-CoV-2 infection | 3 | 0.99 (0.93-1.05) | 0.677 | 0.95 | 0.621 | 0.507 | **0.000** |
| vit. K1 subsignificant | Hospitalized (ver. non-hospitalized) | 3 | 1.06 (0.88-1.28) | 0.565 | 0.50 | 0.779 | 0.697 | **0.000** |
| vit. K1 subsignificant | Hospitalized (ver. population) | 3 | 0.98 (0.87-1.09) | 0.662 | 0.62 | 0.732 | 0.593 | **0.000** |
| vit. K1 subsignificant | Very severe COVID-19 | 3 | 0.93 (0.72-1.19) | 0.546 | 4.42 | 0.110 | 0.349 | 0.084 |

1 - Nominal *p*-value
2 - Insufficient number of SNPs for MR-Egger analysis

**Figure 1.**
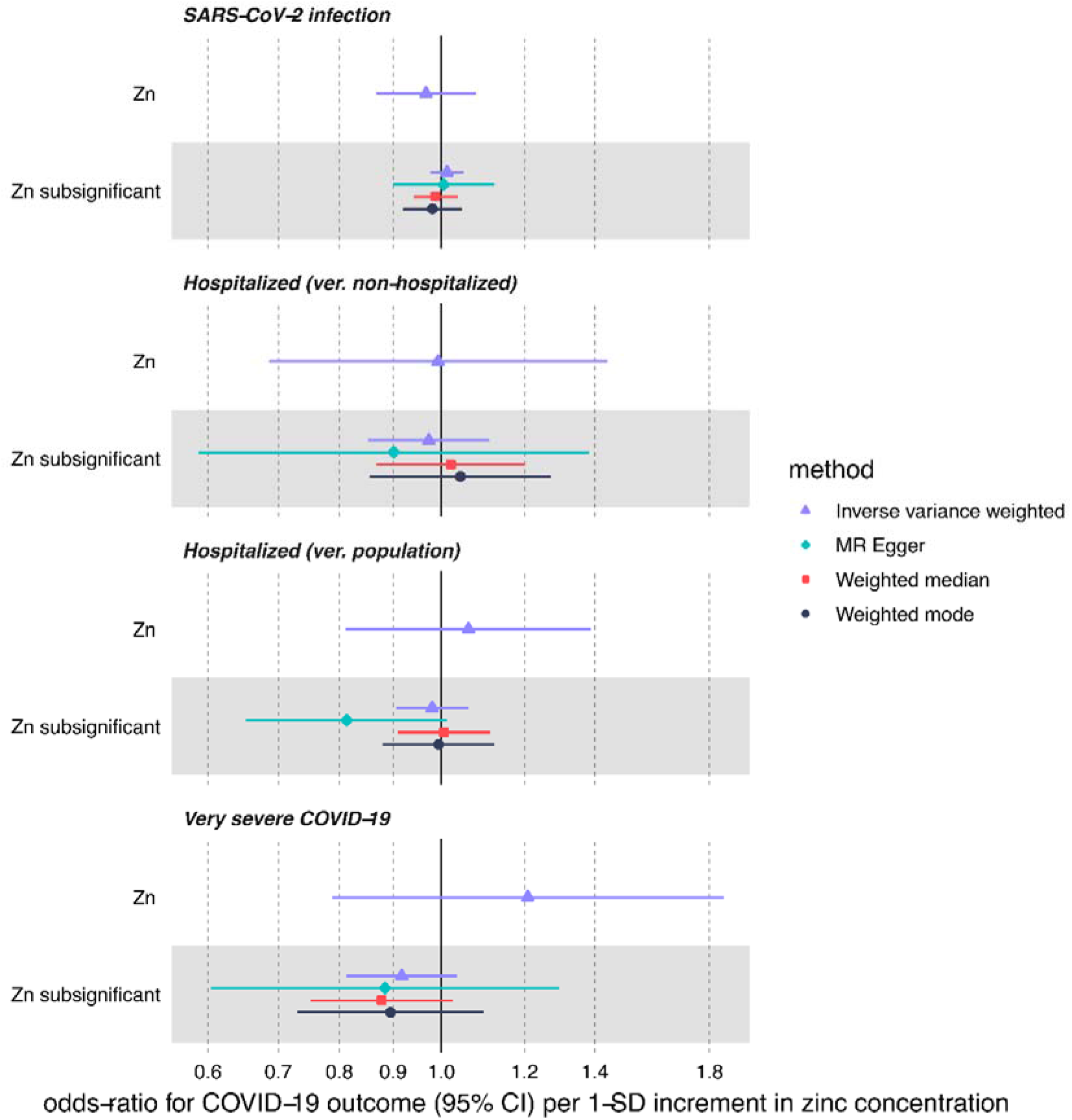
MR estimates for the effect of circulating zinc (Zn) on three COVID-19 outcomes obtained using 4 different statistical methods. We used two sets of zinc instruments: *Zn* refers to instruments with p-values < 5 × 10^−8^ and *subsignificant Zn* refers to instruments with *p*-values < 5 × 10^−5^.

#### Selenium

We found weak causal effect of meta-analysed selenium levels (per 1 SD increment) on SARS-CoV-2 infection, COVID-19 hospitalization and critical illness (Table 1, **Figure 2**). IVW odds ratio of SARS-CoV-2 infection using the instruments in the GWAS meta-analysis of Se blood and toe-nail levels was 1.03 (95% CI: 0.95-1.11, nominal *p*-value=0.5), while in the ALSPAC and QIMR subsignificant (max *p-*value of 5 × 10^−5^) sensitivity analyses we found narrower CI intervals also overlapping 1: OR=0.99 (95% CI: 0.95-1.03, *p*-value=0.70) and 1.00 (0.97-1.03, *p*-value=0.97), accordingly. Similar results were found moving onto protection against hospitalization (ver. population) using the main (OR=0.98; 95% CI: 0.87-1.10, *p*-value=0.71), ALSPAC (OR=1.03, 95% CI: 0.95-1.12, *p-*value=0.45) and QIMR (OR=1.06, 95% CI: 1.00-1.12, *p*-value=0.03) subsignificant sets of instruments. The nominally significant increased risk of hospitalization with COVID-19 in the QIMR cohort did not survive multiple-correction testing. Similar estimates were found in the hospitalized COVID-19 (ver non-hospitalized) comparison, however for that outcome the point-estimate ORs were < 1 in the QIMR cohort using the sensitivity methods: MR-Egger, weighted median and mode (Figure 2, Supplementary Table 8). Finally, increased genetically predicted selenium levels did not causally associate with COVID-19 severity using the meta-analysis (OR=0.99, 95% CI: 0.83-1.17, *p*-value=0.86), ALSPAC (OR=1.06; 95% CI: 0.94-1.19, *p*-value=0.37) or QIMR (OR=1.07; 95% CI: 0.99-1.16, *p*-value=0.07) subsignificant instruments.

**Figure 2.**
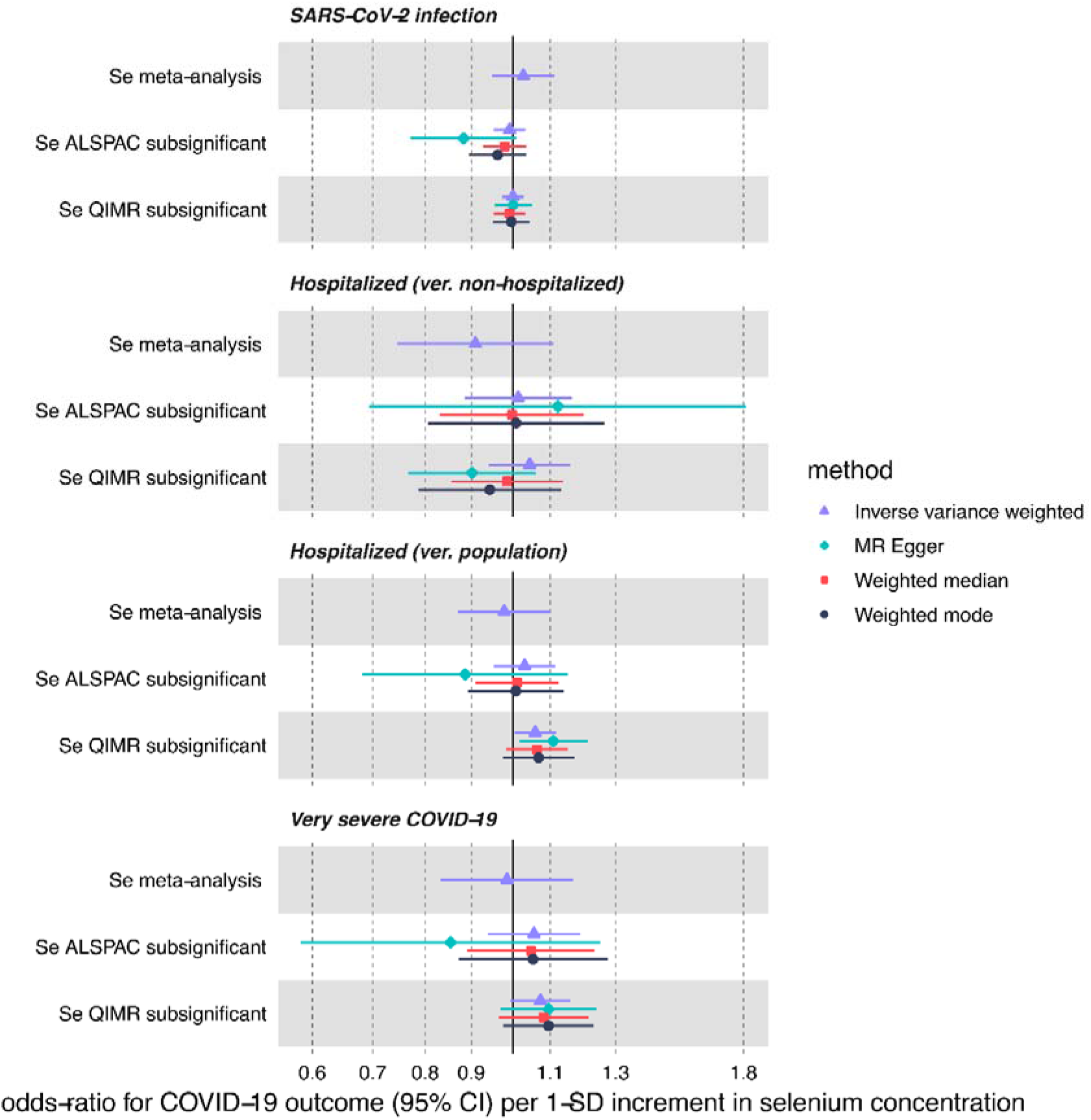
MR estimates for the effect of circulating selenium (Se) on three COVID-19 outcomes obtained using 4 different statistical methods. We used three sets of selenium instruments: Se refers to instruments with *p*-values < 5 × 10^−8^ and *subsignificant Se* refers to instruments with *p*-values < 5 × 10^−5^ in the ALSPAC and QIMR cohorts.

#### Copper

Limited evidence for predicted increase in the risk of SARS-Cov-2 infection per 1 SD increment in circulating copper levels (IVW OR=1.07, 95% CI: 1.00-1.14, nominal *p*-value=0.06) was attenuated (Table 1, **Figure 3**) using the sensitivity subsignificant (max *p-* value of 5 × 10^−5^) instrument set (OR= 1.01; 95% CI: 0.96-1.07, *p*-value=0.66). For the two other outcomes, we found non-significant > 1 OR point estimates in the main analysis (OR= 1.07, 95% CI: 0.88-1.29, *p*-value=0.49 for hospitalized ver. population and OR= 1.13; 95% CI: 0.82-1.55, *p*-value=0.47 for very severe COVID-19) which were flipped < 1 in the subsignificant instrument analysis (OR= 0.99, 95% CI: 0.91-1.08, *p-*value=0.82 and OR= 0.94, 95% CI: 0.83-1.07, *p*-value=0.33 respectively) using IVW method as well as MR-Egger and weighted median (Figure 3, Supplementary Table 8).

**Figure 3.**
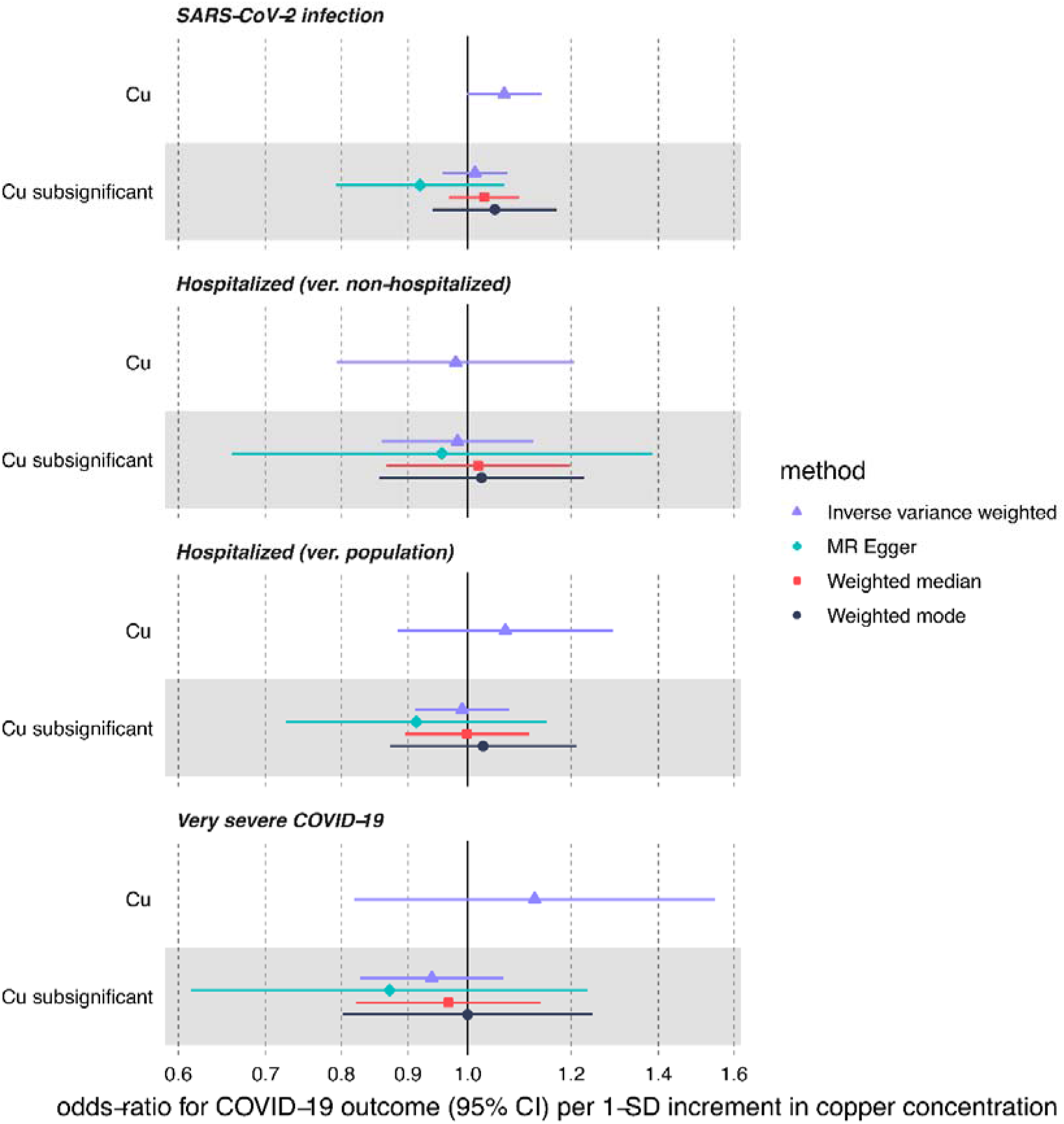
MR estimates for the effect of circulating copper (Cu) on three COVID-19 outcomes obtained using 4 different statistical methods. We used two sets of copper instruments: *Cu* refers to instruments with *p*-values < 5 × 10^−8^ and *subsignificant Cu* refers to instruments with *p*-values < 5 × 10^−5^.

#### Vitamin K_1_

Using a limited set of 3 vitamin K_1_ genome-wide subsignificant (max *p-*value of 5 × 10^−5^) instruments, we were not able to detect any strong effect of genetically-predicted circulating vitamin K_1_ on COVID-19 outcomes (Table 1, **Figure 4**). In the IVW analysis, vitamin K_1_ increment (per natural log transformed nmol/L) associated with OR of 0.99 (95% CI: 0.93-1.05, *p*-value=0.68) for SARS-CoV-2 infection. Next, both hospitalization (ver. population) and very severe COVID-19 showed lower OR on increased vitamin K_1_ exposure (OR=0.98, 95% CI: 0.87-1.09, *p*-value=0.66 and OR=0.93, 95% CI: 0.72-1.19, *p*-value=0.55, respectively) but the CI comfortably overlapped OR either side of 1. The confidence intervals for our sensitivity methods (MR-Egger, weighted median and mode) overlapped the IVW CI, although OR point estimates sometimes differed in terms of direction of effect.

**Figure 4.**
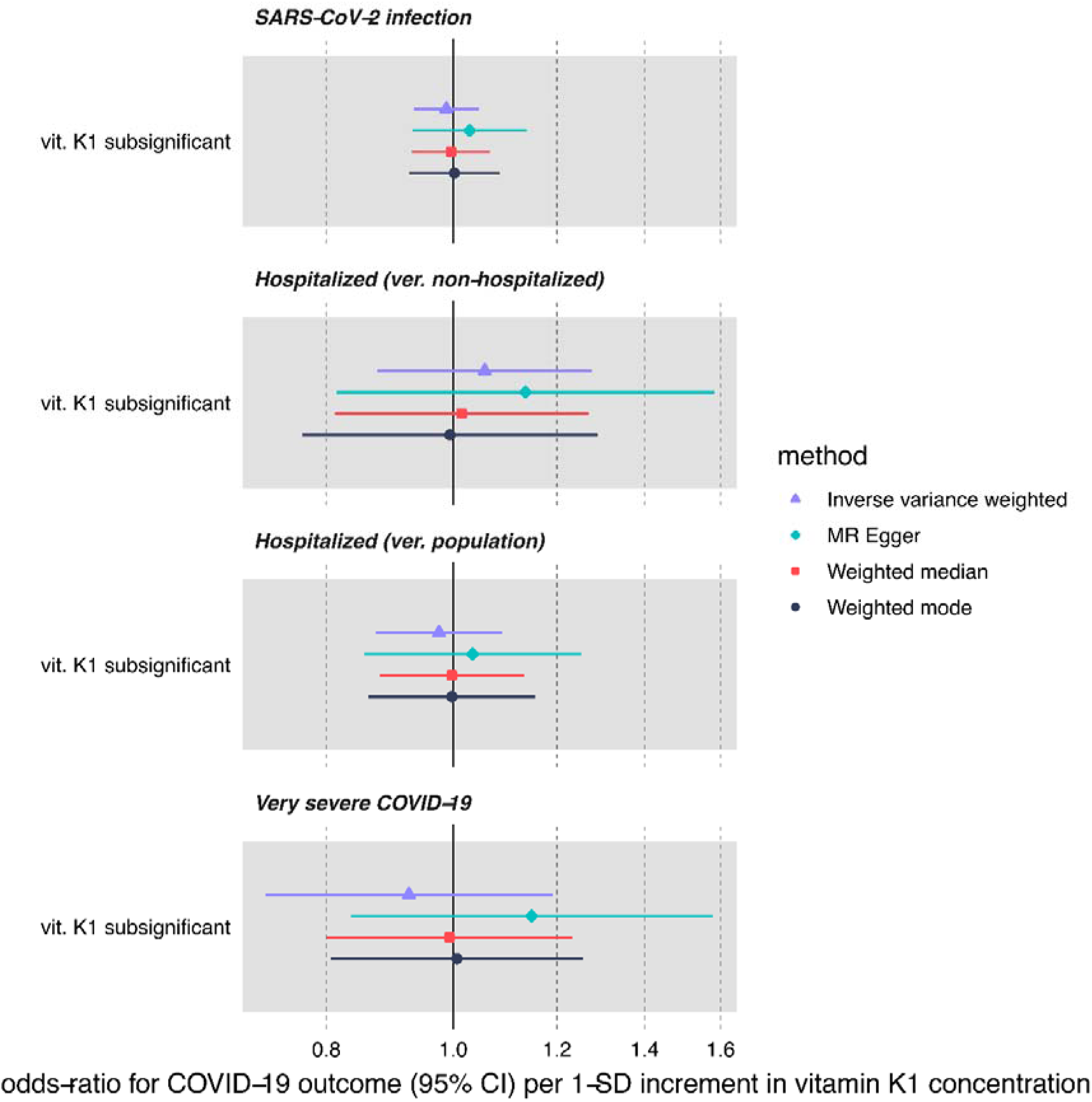
MR estimates for the effect of circulating vitamin K_1_ on three COVID-19 outcomes obtained using 4 different statistical methods. We only had access to instruments with *subsignificant p*-values < 5 × 10.

#### Pleiotropic bias

We detected no significant heterogeneity of effect between our variants using Cochrane’s Q and pleiotropy using MR-Egger intercept in the zinc, selenium and copper MR analysis (Table 1). However, for vitamin K_1_ we did find directional pleiotropy using MR-Egger, with the intercept differing significantly from 0. Therefore, causal estimates using this set of instruments are biased and need to be interpreted cautiously.

## Discussion

Our analyses attempted to elucidate the potential of zinc, selenium, copper and vitamin K_1_ in prophylaxis and treatment of COVID-19 using MR techniques. However, we found little evidence of causal association of genetically predicted micronutrient concentration on COVID-19 outcomes.

Zinc’s many antiviral, immunomodulatory and anti-inflammatory functions have generated a lot of interest for its potential use in COVID-19 management(8,13,49,50,77). Consequently, over 50 RCTs including zinc, albeit typically used as adjuvant treatment or prophylactic, have commenced (ClinicalTrials.Gov, September 2021). One of the first to publish results, The COVID A to Z trial, which tested the direct effect of high-dose zinc supplementation, found no reduction in symptom duration in outpatients and was terminated early(78). In terms of prophylaxis, no beneficial effect of zinc supplementation was reported among 370 000 British users of COVID-19 Symptom Study app(79). However, small observational studies reported lower serum Zn levels as a predictor of illness severity(80–83). As a caveat, all the real life data and MR analyses do not include application of ionophores which may be necessary for zinc’s antiviral inhibition(14,15).

Moreover, a number of observational studies established correlation between low serum selenium levels and COVID-19 severity and mortality(81,84–86). An ecological study found a significant positive association between hair selenium concentration and COVID-19 recovery rate in different provinces of China(87) which was subsequently replicated using local soil selenium concentrations as the predictor variable(88).

In general, one way to partially reconcile our findings for selenium and zinc with general micronutrient deficit in hospitalized or severely ill COVID-19 patients found in small studies comes from previous observations in critically ill individuals. There, the initial hypozincemia and hyposelenemia is thought to stem from disease-driven inflammatory process (acute phase response), is found chiefly in plasma (but not e.g. erythrocytes)(13,89,90) and recovers over time in survivors. However, this does not mean that pre-existing deficiencies will not have a compounded negative impact at this stage(50).

A recently published study suggested that Cu status is correlated with survival status of COVID-19 patients(32). In contrast, another study from Skalny et al.(86) reported that plasma copper levels and Cu/Zn ratio increased in more severe disease. This could reflect the fact that copper and zinc are antagonistically absorbed(91) and high serum Cu/Zn ratio is a marker for infection, as zinc gets redistributed to liver in the acute phase of the infection(92).

Despite evidence of the role of coagulation modulation by vitamin K in COVID-19 severity and poorer outcomes among hospitalized COVID-19 patients with lower vitamin K status(42,43), we did not detect any effect of circulating phylloquinone on very severe COVID-19 or other outcomes.

We carried out multiple sensitivity analyses, involving different methods (e.g. MR-Egger) and instrument selection, which revealed consistent results. In general, inclusion of pleiotropic variants in MR is likely to skew the results away from the null, so it is reassuring that we find no effects also in the analyses involving variants selected at a more liberal subsignificant *p*-value threshold(76) and in vitamin K_1_ analyses showing directional pleiotropic bias.

MR studies have confirmed RCT findings for many known risk factors, such as blood pressure and low-density lipoproteins(93). MR has also indicated no causal effect of high-density lipoproteins and C-reactive protein(94,95) on cardiovascular disease, which, had it been reported earlier, could have saved a lot of effort and expense developing failed drugs. MR evidence can therefore deliver considerable insight about the prospect of a therapeutic. Here, current MR analyses do not support causal pathway between Zn, Se, Cu, vitamin K_1_ blood levels and COVID-19 outcomes.

Thanks to MR framework’s use of genetic instrumental variables, the possibility of confounding and reverse causality was limited. Lately, some concern regarding the ubiquity of collider bias in epidemiological investigations of COVID-19 was voiced(96). However, the use of general population control without known COVID-19 infection in the outcome GWAS serves to decrease the likelihood of collider bias emerging, while not biasing effect size estimates in GWAS sensitivity analyses. We also included a hospitalized versus non-hospitalized GWAS outcome but even in these analyses at a higher risk of collider bias, the results broadly agreed with those from hospitalized versus population outcome. However, ascertainment bias in the GWAS due to differential case reporting and SARS-CoV-2 exposure levels by e.g. socio-economic status remains difficult to account for.

Another potential form of bias affecting MR is population stratification. Since we included only European-ancestry samples in all our analyses, we limit the potential of this bias to skew the results. However, there are no strong biological reasons why our conclusions should not be generalizable to other ancestries.

The main limitation of our study is relatively low power to detect modest effects of micronutrient levels on COVID-19 hospitalization and severity due to few reliable genetic instruments available for micronutrients of interest and limited number of cases in the COVID-19 GWAS. This could be improved in the future as better-powered GWAS for both the exposures (particularly for vitamin K, where no instruments reached genome-wide significance) and outcome become available. Since we used the same GWAS for instrument discovery and effect estimate, our analysis is likely to suffer from “winners’ curse” and increased weak instrument bias that would pull the results towards the null(76), which again could be rectified if new micronutrient GWAS are released.

Another limitation is that MR methods used only model linear effects within normal range of micronutrient concentration, so any potential non-linear U/J-shaped, threshold effects will not be correctly estimated. Individual-level data for both exposure and outcome in the same population sample are required for such an analysis(76). MR also cannot answer the question if specific subgroups, such as micronutrient deficient individuals can benefit from supplementation. Furthermore, phenotypes used in MR analysis typically correspond to lifelong exposure and small changes in micronutrient concentration, which does not exactly mirror intensive, high-dose clinical interventions.

In conclusion, we found little evidence of an effect of genetically predicted zinc, selenium, copper or vitamin K_1_ levels towards preventing infection with SARS-CoV-2, and disease progression: hospitalization or developing very severe COVID-19. Similar MR findings were obtained for two other promising micronutrients: vitamin C(97) and vitamin D(98,99), suggesting that utility of dietary supplementation for general population in the COVID-19 pandemic may be limited.

## Data Availability

All exposure data described in the article is provided in the supplementary tables. All outcome data is available from COVID-19 HGI website: https://www.covid19hg.org/results/r5/ Code for statistical analyses is available on: https://github.com/marynias/covid19

## Statement of authors’ contributions to manuscript

MKS and TRG designed research, MKS analyzed data, MKS and TRG wrote the paper, MKS had primary responsibility for final content. All authors have read and approved the manuscript.

## Sources of support for the work

The supporting source had no involvement in drafting or restrictions regarding publication.

## Online Supporting Material

Supplementary Methods

Supplementary Tables 1-8

## List of abbreviations

GWAS: Genome wide association study
MR: Mendelian Randomization
RCT: Randomized control trial

## Data sharing

All exposure data described in the article is provided in the supplementary tables. All outcome data is available from COVID-19 HGI website: https://www.covid19hg.org/results/r5/. Code for statistical analyses is available on: https://github.com/marynias/covid19

