## Supplementary Methods for "The effect of circulating zinc, selenium, copper and vitamin K_1_ on COVID-19 outcomes: a Mendelian randomization study"

**Vitamin K_1_ genetic instruments**

We removed 4 vitamin K_1_ SNPs from downstream processing. First was the exceptionally pleiotropic (associated with 74 traits in GWAS Catalog and 96 in PhenoScanner) and confounded (p-value close to 1 in 2 of alternative GWAS models) rs964184. Secondly, we excluded rs2108622 (along with rs12609820 in the same clump) which was also highly significantly associated with vitamin E concentration phenotypes. Lastly, we had to discard rs2192574 as it was unavailable in the outcome dataset and no proxy could be found.

**PhenoScanner results**

Out of known risk-factors for COVID-19 with evidence from previous MR analyses(1) – BMI for hospitalization and infection, smoking for hospitalization, height and low red blood cells (RBC) for infection, we found overlap with adiposity, height and RBC traits. One SNP (rs2769264, Cu) was associated with adiposity traits, while 5 with height (rs921943, rs10944 – Se; rs248381, rs17823744, rs2163813 – Se sensitivity analysis). We did not remove the Cu instrument (rs2769264), as it is the strongest and one of only 2 instruments for Cu nutrition, but we note that the Cu associations are by far the strongest at the locus (*p*-value = 2.63 x 10^-20^) and adiposity related traits are only nominally associated (*p-*value > 1 x 10^-7^). Furthermore, copper is known to play an essential role in fat catabolism(2,3) and so BMI is likely a case of vertical pleiotropy, downstream from copper exposure, which does not violate MR assumptions. We were also cautious with SNPs related to height, as height is a famously highly complex polygenic trait(4), and higher COVID-19 reported infection rate in tall individuals currently lacks a plausible biological mechanism. It is causally not clear whether the impact of red blood cell number on COVID-19 infection is not possibly related to their micronutrient content of interest here, so SNPs associated with RBC phenotypes (rs1175550 – Cu, rs2120019 – Zn, rs1532423 – Zn) were also retained. Again, for all those instruments, stronger association was found for micronutrient content relative to RBC phenotypes, except for rs1175550, despite pronounced difference in respective GWAS power: *n*=2,603 for Zn and Cu, *n*= 173,480 for RBC phenotypes; these SNPs were also associated with other blood cell phenotypes.
